# A Retrospective Study of Lung/Bronchus Cancer Mortality Rates Before and After Enactment of the Affordable Care Act in Mississippi and New York

**DOI:** 10.1101/2025.07.23.25331857

**Authors:** Alessandro Drudi, Josephine Chan, Bei Peng, Darwin Jean, Tristan Singh, Mark Richman

**Affiliations:** Northwell Health Long Island Jewish Medical Center, Department of Emergency Medicine, 270-05 76th Ave., New Hyde Park, NY 11004.

## Abstract

**Introduction:** The Affordable Care Act (ACA), whose major provisions were Medicaid expansion, state insurance exchanges, and allowing dependents to remain on their parents’ insurance until age 26, was implemented fully in 2014. Increased access to care might have improved access to lung/bronchus cancer diagnosis, treatment, and outcomes. We hypothesized differential changes in lung/bronchus mortality among Mississippi (which did not expand Medicaid) and New York (that did expand Medicaid).

**Methods:** We utilized the CDC Wonder database to compare lung/bronchus cancer mortality rates in Mississippi, New York, and the United States as a whole, comparing such rates 5 years prior (2009-2013) to 5 years after (2015-2019) ACA enactment. Statistical significance was deemed at p <0.05.

**Results:** In the 5 years prior to ACA implementation (2009-2013), there was no statistically-significant difference between Mississippi, New York, or the total U.S. in the percent of population dying from lung/bronchus cancer (p >0.7). In the 5 years following ACA implementation (2015-2019), there was a statistically-significant decrease between Mississippi, New York, and the total U.S. in the percent of population dying from lung/bronchus cancer (p <0.002). Comparing the years 2009-2013 (5 years prior to ACA) and the years 2015-2019 (5 years after ACA), there was a statistically-significant difference in the decrease in percent of population dying from lung and bronchus cancers in New York, Mississippi, and the total United States (p <0.002), with New York having the greatest percent decline. In New York, the percentage of mortalities from lung and bronchus cancers decreased from 0.004691% to 0.004088% (p <0.0001). In Mississippi, the percentage of mortalities from lung and bronchus cancers decreased from 0.006537% to 0.006282% (p = 0.0061). In the total US, the percentage of mortalities from lung and bronchus cancers decreased from 0.05097% to 0.0473% (p <0.0001).

**Conclusion:** State-level participation in the ACA’s Medicaid expansion was associated with disproportionate improvement in lung/bronchus mortality compared with non-participation and with the U.S. in total.

## Introduction

The Affordable Care Act, colloquially known as Obamacare, was implemented fully in 2014 and resulted in “gains in health insurance coverage for 20 million adults through early 2016”.^1^ The major provisions that resulted in this increase were Medicaid expansion (accomplished by increasing the upper level of poverty qualifying for Medicaid), state insurance exchanges, and allowing dependents to remain on their parents’ insurance until age 26.

Considering this large increase in the number of insured individuals who had been previously uninsured, it can be expected that there were broad changes in health care visits, processes, procedures, and outcomes. Many studies have already been published regarding how the ACA has affected outcomes. For example, a pilot study in Multnomah County, Oregon found a significant reduction in out-of-hospital cardiac arrest (OHCA) incidence associated with Medicaid Expansion in the state.^2^ Another potential change is an increase in cancer diagnoses and access to treatment. An increase in insurance availability would potentially allow previously-undiagnosed cancers to be detected and treated. A particular type of cancer that is notable primarily due to it having a high incidence rate and prevalence is lung/bronchus cancer. Lung/bronchus cancer is the second-most diagnosed cancer in both men and women (behind prostate cancer in men and breast cancer in women). Lung/bronchus cancer in the last 5 years has also had a relatively-large improvement in death rates (−3.0% in females and −4.4% in males)^3^.

This study aims to investigate the correlation between ACA implementation and lung/bronchus cancer mortality rate in all age groups for 5 years before ACA implementation and 5 years after ACA implementation. We will compare one state that did (New York) and one that did not (Mississippi) expand Medicaid as part of the ACA. These states also offered different levels of infrastructure, and healthcare availability per capita before the ACA so it will be relevant to adjust for this assistance. In a 2022 CDC study, Mississippi had the highest rate of cancer deaths per 100,000 population at 178.4, and New York had the third lowest rate of cancer deaths per 100,000 population at 122.4.^4^ In 2010, Mississippi’s per capita healthcare spending was $6,554, while in New York it was $8,759. In 2018, Mississippi’s per capita healthcare expenditure was $8,417,^5^ while New York’s was $12,098.^6^

We hypothesize that, even adjusting for differences in pre-ACA healthcare infrastructure, lung/bronchus mortality rates among New York residents would drop greater than among Mississippi residents, on account of ACA adoption.

## Methods

The lung/bronchus cancer mortality rates in Mississippi and New York were compared between 2009 to 2019 (5 years prior to 5 years after enactment of the ACA) were studied in all age groups. The database used to retrieve this data was CDC Wonder, which compiles data from the Centers for Disease Control and Prevention’s National Program of Cancer Registries (NPCR), The National Cancer Institute’s Surveillance, Epidemiology, and End Results (SEER), and the Centers for Disease Control and Prevention’s National Vital Statistics System (NVSS). The number of lung/bronchus cancer deaths in both states, and of the U.S. as a whole (as a control) was compared to the respective state’s (and the U.S.) total population; these comparisons determined a mortality/population percentage. This study observed the changes in lung/bronchus mortality percentages within both states and the total U.S. population between 2009-2019.

For both states and the United States, the number of mortalities from 2009-2013 and 2015-2019 was totalled and divided by the total population during those time frames. This allowed calculation of the mortality/population percent. A <u>chi square test</u> compared the mortality/population percentages for Mississippi, New York, and the U.S. in total during each 5year timeframe (2009-2013 and 2015-2019). A separate chi square test compared the change in mortality/population percentages for Mississippi, New York, and the U.S. in total between the 2009-2013 timeframe and the 2015-2019 timeframe.

Adjustment for pre-ACA infrastructure was attained by considering pre-ACA per capita spending and pre-vs. post-ACA changes in private health insurance coverage, as both of these would have been outside of the influence of the ACA.

Microsoft Excel was used to analyze the percentage change in mortality/population within each state and between different states from 2009 to 2019. Statistical significance was deemed at p <0.05.

This project was reviewed and deemed not to meet the guidelines of being a research project by the Northwell Health Institutional Review Board’s (IRB’s) Human Research Protection Program (HSRD24-0133), thus denoting that formal IRB approval was not necessary for this study.

## Results

In the 5 years prior to ACA implementation (2009-2013), there was no statistically-significant difference between Mississippi, New York, or the total U.S. in the percent of population dying from lung/bronchus cancer (p >0.7) (*Table 1*). In the 5 years following ACA implementation (2015-2019), there was a statistically-significant decrease between Mississippi, New York, and the total U.S. in the percent of population dying from lung/bronchus cancer (p<0.002) (*Table 2*). Comparing the years 2009-2013 (5 years prior to ACA) and the years 2015-2019 (5 years after ACA), there was a statistically-significant difference in the decrease in percent of population dying from lung and bronchus cancers in New York, Mississippi, and the total United States (p <0.002) (*Table 3*), with New York having the greatest percent decline.

**Table 1.**
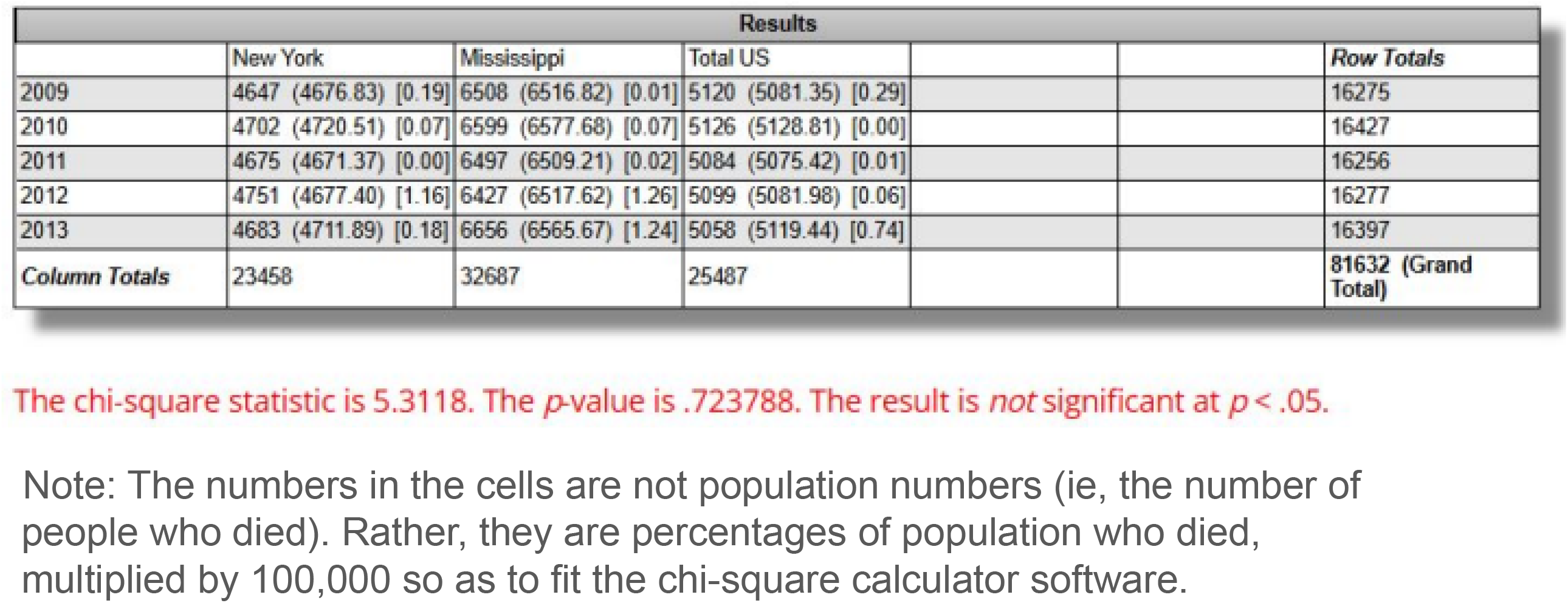

| Results |  |  |  |  |  |  |
| --- | --- | --- | --- | --- | --- | --- |
|  | New York | Mississippi | Total US |  |  | Row Totals |
| 2009 | 4647 (4676.83) [0.19] | 6508 (6516.82) [0.01] | 5120 (5081.35) [0.29] |  |  | 16275 |
| 2010 | 4702 (4720.51) [0.07] | 6599 (6577.68) [0.07] | 5126 (5128.81) [0.00] |  |  | 16427 |
| 2011 | 4675 (4671.37) [0.00] | 6497 (6509.21) [0.02] | 5084 (5075.42) [0.01] |  |  | 16256 |
| 2012 | 4751 (4677.40) [1.16] | 6427 (6517.62) [1.26] | 5099 (5081.98) [0.06] |  |  | 16277 |
| 2013 | 4683 (4711.89) [0.18] | 6656 (6565.67) [1.24] | 5058 (5119.44) [0.74] |  |  | 16397 |
| Column Totals | 23458 | 32687 | 25487 |  |  | 81632 (Grand Total) |

The chi-square statistic is 5.3118. The *p*-value is .723788. The result is *not* significant at *p* < .05.
Note: The numbers in the cells are not population numbers (ie, the number of people who died). Rather, they are percentages of population who died, multiplied by 100,000 so as to fit the chi-square calculator software.

**Table 2.**

| Results |  |  |  |  |  |  |
| --- | --- | --- | --- | --- | --- | --- |
|  | New York | Mississippi | Total US |  |  | Row Totals |
| 2015 | 4413 (4285.40) [3.80] | 6437 (6585.65) [3.36] | 4979 (4957.95) [0.09] |  |  | 15829 |
| 2016 | 4198 (4178.19) [0.09] | 6413 (6420.89) [0.01] | 4822 (4833.91) [0.03] |  |  | 15433 |
| 2017 | 4036 (4017.65) [0.08] | 6080 (6174.18) [1.44] | 4724 (4648.18) [1.24] |  |  | 14840 |
| 2018 | 3990 (4003.03) [0.04] | 6194 (6151.71) [0.29] | 4602 (4631.26) [0.18] |  |  | 14786 |
| 2019 | 3804 (3956.73) [5.90] | 6289 (6080.57) [7.14] | 4522 (4577.70) [0.68] |  |  | 14615 |
| Column Totals | 20441 | 31413 | 23649 |  |  | 75503 (Grand Total) |

The chi-square statistic is 24.3705. The $p$ -value is .001986. The result is significant at $p < .05$ .
Note: The numbers in the cells are not population numbers (ie, the number of people who died). Rather, they are percentages of population who died, multiplied by 100,000 so as to fit the chi-square calculator software.

**Table 3.**

| Results |  |  |  |  |  |  |
| --- | --- | --- | --- | --- | --- | --- |
|  | New York | Mississippi | Total US |  |  | Row Totals |
| 5 Years Prior | 4691 (4560.61) [3.73] | 6537 (6659.35) [2.25] | 5097 (5105.04) [0.01] |  |  | 16325 |
| 5 Years After | 4088 (4218.39) [4.03] | 6282 (6159.65) [2.43] | 4730 (4721.96) [0.01] |  |  | 15100 |
| Column Totals | 8779 | 12819 | 9827 |  |  | 31425 (Grand Total) |

The chi-square statistic is 12.463. The $p$ -value is .001967. The result is significant at $p < .05$ .
Note: The numbers in the cells are not population numbers (ie, the number of people who died). Rather, they are percentages of population who died, multiplied by 100,000 so as to fit the chi-square calculator software.

In New York, the percentage of mortalities from lung and bronchus cancers decreased from 0.004691% to 0.004088% (a decrease of 0.00603%, p <0.0001) (*Table 4*). In Mississippi, the percentage of mortalities from lung and bronchus cancers decreased from 0.006537% to 0.006282% (a decrease of 0.00205%, p = 0.0061) (*Table 5*). In the total US, the percentage of mortalities from lung and bronchus cancers decreased from 0.05097% to 0.0473% (a decrease of 0.00368%, p <0.0001) (*Table 6*). (*Figure 1*).

**Table 4.**

| *Table 4* | Mortalities | Population<br>(2010) | Mortalities/<br>Population | 5-year Average<br>of Mortalities | Total<br>Mortalities | Total<br>Mortalities over<br>Sum of<br>Population |
| --- | --- | --- | --- | --- | --- | --- |
| 2009 | 9,005 | 19,378,102 | 0.04647% | 9,091.20 | 45,456.00 | 0.04691% |
| 2010 | 9,112 | 19,378,102 | 0.04702% |  |  |  |
| 2011 | 9,059 | 19,378,102 | 0.04675% |  |  |  |
| 2012 | 9,206 | 19,378,102 | 0.04751% |  |  |  |
| 2013 | 9,074 | 19,378,102 | 0.04683% |  |  |  |
| 2014 | 8,563 | 19,378,102 | 0.04419% |  |  |  |
| 2015 | 8,551 | 19,378,102 | 0.04413% | 7,922.20 | 39,611.00 | 0.04088% |
| 2016 | 8,135 | 19,378,102 | 0.04198% |  |  |  |
| 2017 | 7,821 | 19,378,102 | 0.04036% |  |  |  |
| 2018 | 7,732 | 19,378,102 | 0.03990% |  |  |  |
| 2019 | 7,372 | 19,378,102 | 0.03804% |  |  |  |
| Lung and Bronchus Cancer Mortalities in New York 5 years prior and after 2014 Affordable Care Act implementation |  |  |  |  |  | Difference: -0.00603% |

**Table 5.**
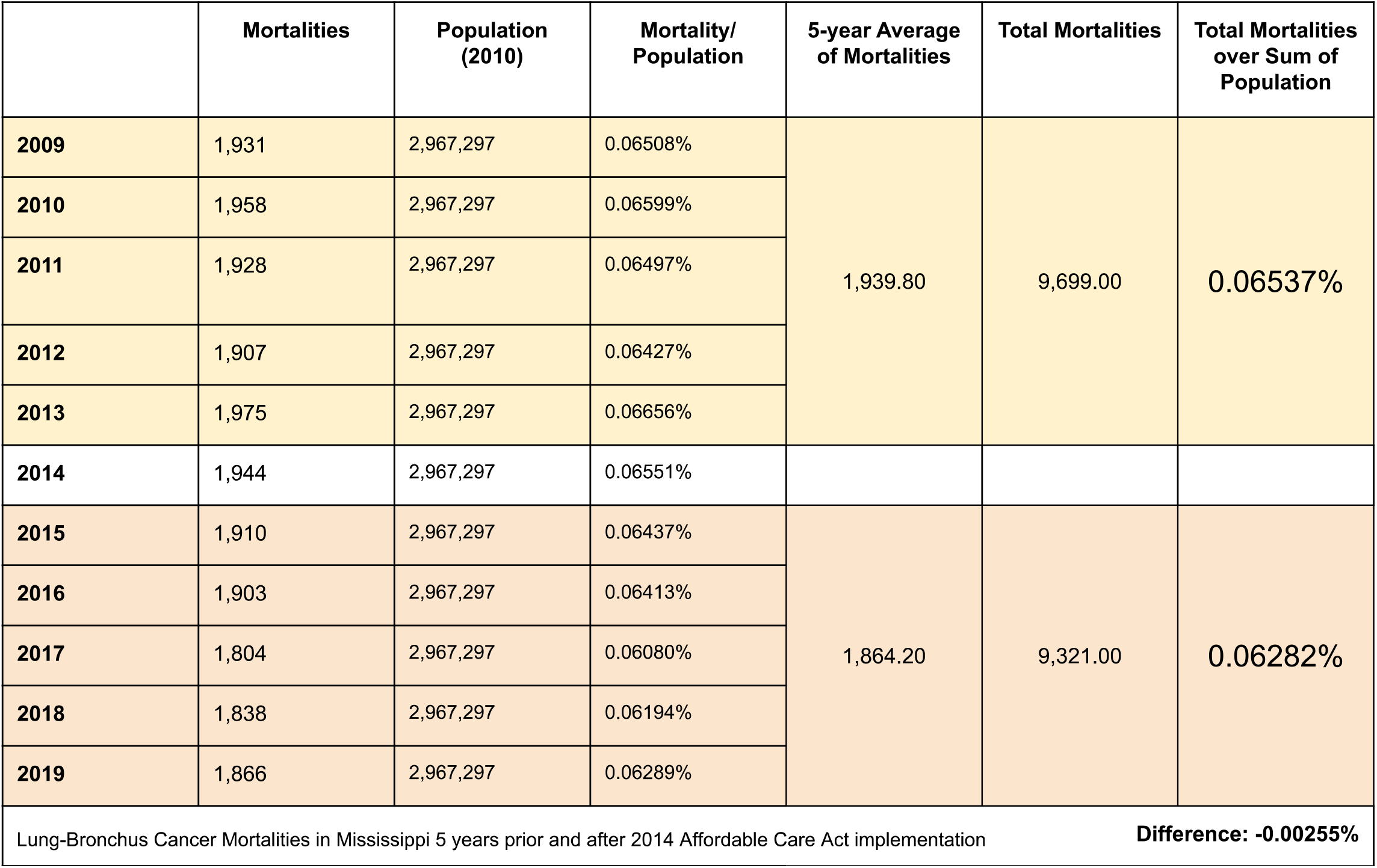

**Table 6.**

| *Table 6* | Mortalities | Population (2010) | Mortality/ Population | 5-year Average of Mortalities | Total Mortalities | Total Mortalities over Sum of Population |
| --- | --- | --- | --- | --- | --- | --- |
| 2009 | 158,081 | 308,745,538 | 0.05120% | 157,376.20 | 786,881.00 | 0.05097% |
| 2010 | 158,248 | 308,745,538 | 0.05126% |  |  |  |
| 2011 | 156,953 | 308,745,538 | 0.05084% |  |  |  |
| 2012 | 157,423 | 308,745,538 | 0.05099% |  |  |  |
| 2013 | 156,176 | 308,745,538 | 0.05058% |  |  |  |
| 2014 | 155,526 | 308,745,538 | 0.05037% |  |  |  |
| 2015 | 153,718 | 308,745,538 | 0.04979% | 146,023.40 | 730,117.00 | 0.04730% |
| 2016 | 148,869 | 308,745,538 | 0.04822% |  |  |  |
| 2017 | 145,849 | 308,745,538 | 0.04724% |  |  |  |
| 2018 | 142,080 | 308,745,538 | 0.04602% |  |  |  |
| 2019 | 139,601 | 308,745,538 | 0.04522% |  |  |  |
| Lung and Bronchus Cancer Mortalities in the United States 5 years prior and after 2014 Affordable Care Act implementation |  |  |  |  |  | Difference: -0.00368% |

**Figure 1.**
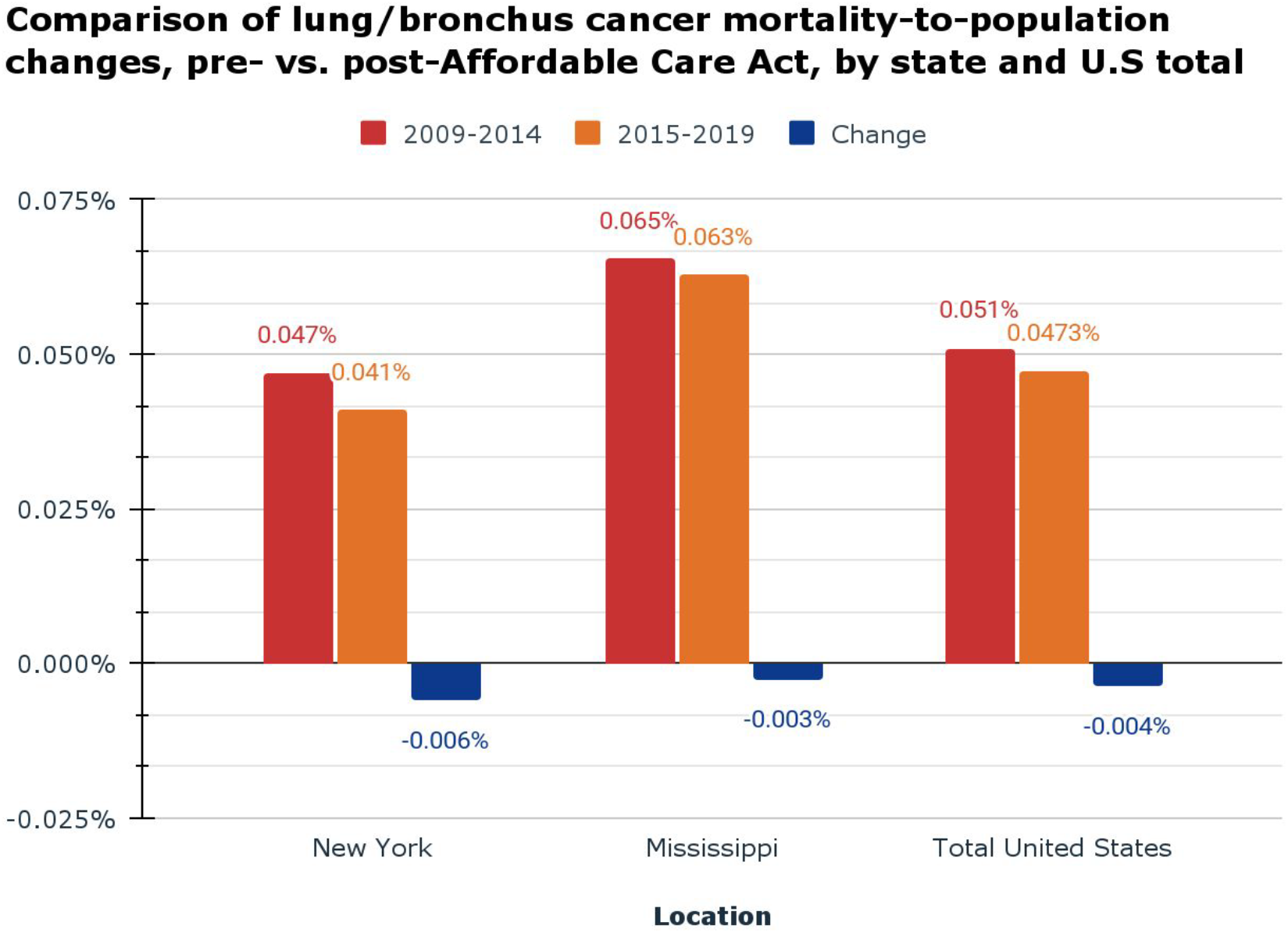

## Discussion

The percentage of population dying from lung and bronchus cancers in Mississippi, New York, and the U.S. in total decreased by a statistically-significant amount after ACA implementation. The magnitude of decline differed between the three venues, being greatest in New York, which participated in the Medicaid expansion.

These results support the possibility that differential changes in lung/bronchus mortality are associated with New York participating in the ACA’s Medicaid expansion program while Mississippi didn’t (and the U.S. in total is a blend of 41 states that did and 9 that didn’t).^7^

Many potential confounding variables might explain these differential decreases in lung/bronchus cancer mortality. These include:

1. The distribution of lung/bronchus cancer mortality by different insurance statuses. Our data did not contain patient insurance, so we were unable to distinguish whether major improvements in lung/bronchus cancer mortality were concentrated among particular insurance groups, such as private insurance vs. Medicare vs. Medicaid. For example, it is possible that most of the improvements in lung/bronchus cancer mortality occurred among patients with private insurance and not among those with Medicaid.
2. The availability by insurance plan of new cancer therapies^8^
3. Varying density of healthcare services (eg, providers,^9^ imaging centers,^10^ hospitals^11^).

This preliminary study will be followed with a multivariate analysis of such confounders.

## Data Availability

All data produced in the present study are available upon reasonable request to the authors

https://wonder.cdc.gov/

